# KNOWLEDGE OF AND PRACTICES AROUND ZOONOTIC DISEASES AMONGST ACTORS IN THE LIVESTOCK TRADE IN THE LAKE VICTORIA CRESCENT ECOSYSTEM IN EAST AFRICA

**DOI:** 10.1101/2022.03.10.22272135

**Authors:** Hamilton Majiwa, Salome A. Bukachi, Dalmas Omia, Eric M. Fèvre

## Abstract

**Background:** Zoonotic diseases pose a direct threat to health and undercut livelihoods in the communities in which they occur. A combination of anthropogenic, animal, and ecosystem activities drives the emergence and re-emergence of zoonotic diseases. Consequently, One Health approaches are necessary to alleviate disease impacts. To be effective, understanding people’s Knowledge, Attitudes, and Practices concerning these disease threats are essential for their prevention, control, and eventual elimination. Livestock traders interact closely with livestock, which puts these traders at high risk of infection and creates conditions by which they may spread zoonotic pathogens through the animals they trade. It is, thus, essential to examine practices among actors involved in livestock trade to understand how well to mitigate these risks in a non-pastoral production system.

**Methods:** Busia County was selected for this study because it is a predominantly crop-producing area, with cross-border (between Kenya and Uganda) trade in livestock. A qualitative study was conducted among the actors in the livestock trade on their knowledge, attitudes, and practices that may contribute to the spread, control, and prevention of zoonotic disease transmission. A thematic analysis framework was used to categorize and synthesize data from In-depth interviews (IDIs) and the Key informant interviews(KIIs).

**Results:** Whereas participants could list the signs of zoonotic diseases, they could not identify these diseases by name, which, demonstrates insufficient knowledge of zoonosis. Brucellosis, Foot and Mouth Disease (FMD), and Anthrax were broadly mentioned as diseases of importance by many actors this shows that they are the common livestock diseases in the area. The actors identified sick animals by checking for dropped ears, mass mucus production; diarrhea; bloody urinal discharge; and general animal activity levels, an animal that is not actively eating or walking will be a sign of sickness. To manage the spread of these diseases, they wash their animals, isolate sick animals from the rest of the stock; vaccinate their animals against certain diseases as preventive practices, they also seek help from animal health professionals for the sick animals as curative practices. The practices of skinning dead animals before burying them and the consumption of dead carcasses risk the increase of zoonotic disease transmission. These practices are drawn from cultural values and beliefs about the curses and potential loss of entire stock if livestock is unceremoniously disposed of upon death, irrespective of the cause.

**Conclusions and recommendations:** Livestock actors have agency in the prevention and elimination of zoonotic diseases, hence, they need to be involved when developing intervention programs and policies for the extension services. Training these actors as a continuum of animal health workers blends lay and professional knowledge, which alongside their intense contact with large numbers of animals, becomes a critical disease surveillance tool this may also play a role in supporting state actors in disease surveillance and response. There is an urgent need to increase awareness of zoonoses within livestock keepers and traders, these campaigns, need should deploy multi-disciplinary teams with an understanding of human health, animal health, and social scientists so that the risky but deeply rooted traditional practices like skinning and eating dead animals can be minimized.

## Introduction

Any infectious disease potentially transmissible from animals, both wild and domestic, to humans is defined as a zoonotic disease (1). The diseases that infect humans from animals are caused by bacterial, viral, parasitic, or fungal infection and spread to humans through bites, scratches, vectors, or ingestion. Zoonotic diseases are categorized according to their route of transmission, namely; vector-borne or foodborne, pathogen types, such as microparasites, viruses, bacteria, protozoa, worms, ticks, or fleas, or degree of person-to-person transmissibility (2).

Humans live in close relationships with domesticated animals or those chosen as pets. These animals carry pathogens that are transmissible to humans and can be harmful to health (3). Zoonotic diseases pose problems to global health. A World Health Organization (WHO) Report 2006 (4) lists rabies, Human African Trypanosomiasis (HAT), brucellosis, bovine tuberculosis, cysticercosis, echinococcosis, and anthrax as the endemic diseases of concern.. These diseases pose health burdens in addition to negative social and economic effects on communities. The infected individual becomes unproductive, and close relatives spend money providing care and treatment. Time and money spent searching for a cure may put a severe drain on the family resource (5). The varying public health burden and social-economic impact of zoonotic diseases across time and geographical settings make prioritizing their prevention and control important (6). The emergence of zoonoses, both recent and historical, can be considered as a logical consequence of both pathogen and human ecology and pathogen evolution, as microbes exploit new niches and adapt to new hosts(7). Access to these new niches is mediated by human action in most cases, including changes in land use, extraction of natural resources, animal production systems, modern transportation, antimicrobial drug use, and global trade (7).

Although underlying ecological principles that shape how these pathogens survive and change have remained similar, people have changed the environment in which these principles operate. Domestication of animals, clearing of land for farming and grazing, and hunting wildlife in new habitats have resulted in zoonotic human infection with microorganisms that cause diseases (8). As human societies have developed, each era of the livestock revolution has presented new health challenges and new opportunities for the emergence of zoonotic pathogens (9). The appearance and re-appearance of many diseases, including zoonotic diseases, has been driven by the changing and increasing interconnection among humans, animals, and the ecosystem over time. Cultural changes as a result of the rise in population, economic and technical developments, and intensification of farming (10) have created more intense interaction between humans and livestock; the increased risk of disease emergence and the possibility for pervasiveness have been linked to increased regional trade and travel, swelling human and livestock populations, and evolving cultures in recent times (11). This warrants the need for continuous surveillance and disease monitoring and an understanding of the cultural epidemiology of zoonotic diseases.

Social aspects of epidemiology include people’s cultures and traditions, norms, knowledge, attitudes, and practices, contributing to the spread, control, and elimination of various zoonotic diseases (12). Studies have shown that knowledge of reservoirs of zoonoses and how they are transmitted to humans has enabled early detection and reporting and their control (13). Evidence shows that people’s perceptions about disease risks such as transmission and health consequences influence their attitudes and health-seeking actions and behaviors towards the diseases concerned (14). Knowledge or awareness about particular diseases vary across individuals and communities

The livestock trade system is a complicated chain with producers, traders, and numerous other market participants all referred to here as actors in the livestock trade. Animals travel from homes through different markets and trade routes to the last consumer or terminal market (15). Numerous informal chains with independent livestock and meat traders play a critical role in the livestock trade (16).

The flow diagram below (Fig 1.0) shows how various actors in the livestock trade are connected

**Figure 1.0:**
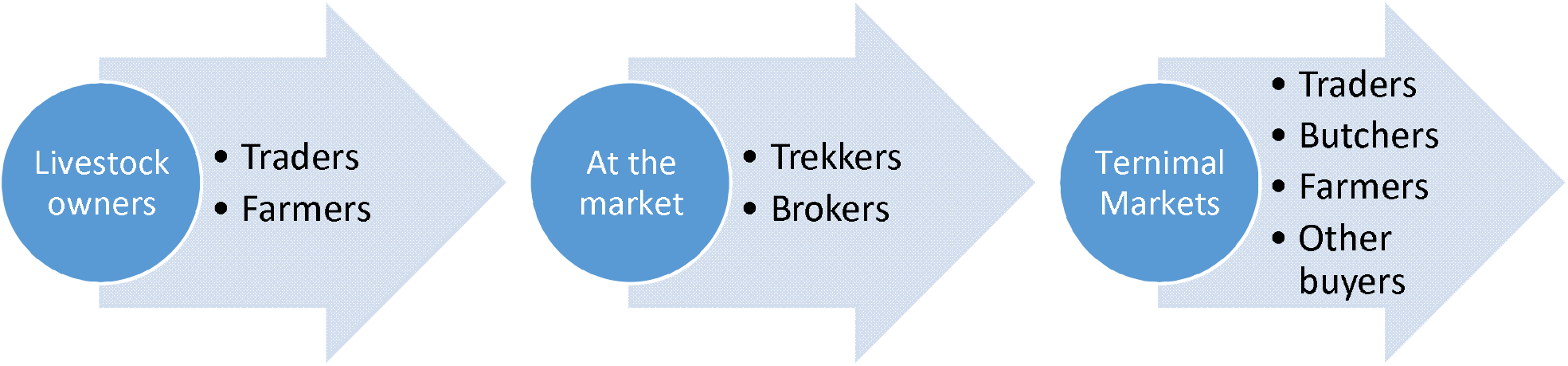
Different actors in livestock trade

The actors include the livestock keepers, who rear the livestock and make the decision to sell them. Brokers are mainly involved in connecting potential sellers to buyers. There are traders whose business is to buy and sell the livestock. There are also animal trekkers whose role is to walk the livestock from the homes to the markets or across markets. The East African Community (EAC) partner states share a similar disease profile (17), their borders are also highly porous to informal animal and human movement, which is common in countries where animal production is not intensive(18). Because of the ease of movement, zoonotic diseases can spread across international borders, hence, the Lake Victoria Crescent ecosystem, which has a dense human population, cross border trade and intensifying farming was chosen for this study. The Lake Victoria Crescent forms the right environment for strategic zoonotic disease control programs through public health awareness campaigns among local and foreign livestock traders. The area is occupied by non-pastoralist communities, which departs from the traditional focus on pastoralism when considering animal movement, yet, it is a critical area in relation to livestock and zoonotic diseases. The study sought to bring out the knowledge and practices on zoonotic diseases in non–pastoral areas and more so, among actors in livestock trade in the Lake Victoria Crescent ecosystem.

## Methods

### Study Area

The study was conducted in Busia County, Western Kenya (Figure 1.1). Busia County is located in the western part of Kenya and broadly represents the Lake Victoria Crescent Ecosystem (19), where several zoonotic infections are co-endemic (20). It borders Uganda with two border crossing points at Busia and Malaba towns. It is one of the four counties comprising the Western Kenya region and situated at the extreme western border of the country

**Figure 1.1:**
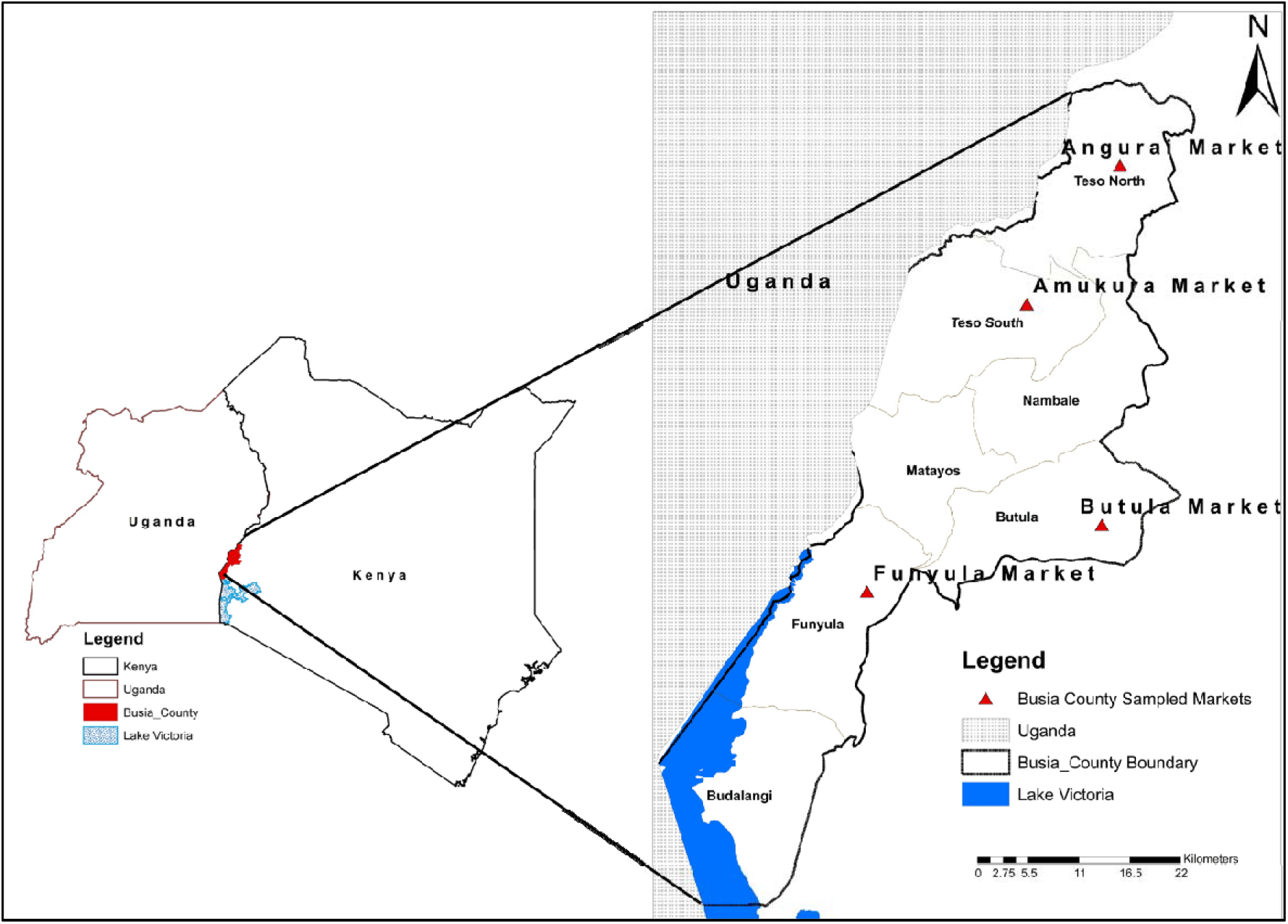
Map of the markets in Busia County in Western Kenya region

### Research design

This exploratory study used in-depth interviews, key informant interviews, and unstructured observation to understand the knowledge, attitudes, and practices of actors involved in the livestock trade. The study began with in-depth interviews with livestock traders and trekkers on their knowledge, attitudes, and practices (KAP) in relation to zoonotic diseases. This was followed by key informant interviews with the market chairman, chairman of the traders, chairman of the trekkers, officers in charge of animal health at the market, and the market masters who are County officials in the market. The in-depth interviews teased out the actors’ KAP on zoonotic diseases; the key informant interviews then followed this. The key informant interviews aimed to provide more insight into the actors’ knowledge of the diseases, the perceptions of the diseases at the community level, and some of the practices that may contribute to spreading or controlling zoonotic diseases. The audio data was transcribed once the qualitative data from the in-depth interviews and key informant interviews were obtained. Where the interviews were not conducted in English the audio data was translated and transcribed. The transcription was done verbatim. Thematic framework analysis was used to systematically categorize and synthesize the qualitative data the interviews generated. All transcripts were read and reread for salient themes

### Sample and Sampling procedure

Purposive sampling (21) was employed to identify 30 actors in the livestock traders. The traders and trekkers were identified with the help of their chairman. Informants who included animal health officers, markets officials, and county government officials were sampled purposefully for key informant interviews.

### Data collection methods

In-depth interviews were conducted with thirty informants, the interviews lasted between 25 to 30 minutes. The interviews were conducted in the markets but away from the crowds to enable the informants to continue their operation without much interference. Curious onlookers were explained to that only the responses of the selected respondent were needed at that time The interviews were audio-recorded with permission from the informants.

Key informant interviews were also carried out with nine key informants The interviews were also audio-recorded with permission from the informants.

Unstructured observations were made on the interactions among the livestock actors and the livestock within the market space.

### Ethical considerations

Participants gave informed, written consent before participation The study was approved by the International Livestock Research Institute Institutional Research Ethics Committee in Kenya (ILRI-IREC2017-08), which is registered and accredited by the National Commission for Science, Technology and Innovation in Kenya, and approved by the Federal wide Assurance for the Protection of Human Subjects in the USA. A research permit was obtained from the National Commission for Science, Technology, and Innovation (NACOSTI) License No: NACOSTI/P/19/2547.

## Results

### Profile of the respondents

This study comprised thirty informants with different roles interviewed using in-depth interviews, key informant interviews, and observations. There were only male trekkers aged 18-50 in the study. The study found that a majority of them had not completed primary school education. Most of the traders were aged above 30 years with the oldest being 60. As opposed to the trekkers, who were all male, the study found three female traders. All trekkers and a majority of traders identified themselves as Christians, while a paltry (<10 %) identified themselves as Muslims. The trekkers charge about 2 USD per animal that they move. The average market-day earnings for the trekkers were established to be $10 relative to $50 by the traders.

Like in a previous study (22), age, education level, religion, and sex had no significant effect on actors’ knowledge level and awareness toward zoonotic diseases as the responses given did not differ much depending on these variables. However, religious affiliation affected their practices like some do not eat animals that have not been slaughtered because this goes against their religious teaching or because they are born-again Christians and also there were Muslims who do not interact with pigs in any way. The ethnicity of the traders and trekkers was Abaluhya (80%) Ateso (15%) and the Luo(5%). The different ethnic groups have varied beliefs and practices, as shown in the findings where the Abaluhya and Ateso don’t believe in throwing away meat or burying dead animals without skinning them. Some Luos also don’t believe in throwing away meat from dead animals and would rather share it with people if they won’t eat it themselves.

The study found that many traders are introduced into the trade through an apprenticeship by their relatives or friends. Some were also driven to the trade by circumstances, need to earn a living. Starting the practice for trekkers seems to require low capital investment in terms of financial capital but more in terms of social capital, they need to be trusted, reliable and popular to get business from the traders as reflected in these excerpts:

> “*You need to be known to the traders and the buyers and to be trusted because someone cannot give you their animal if they don’t trust you and know where you come from”* (**trekke**r)
>
> *“For you to start as a trekker, you don’t need capital; you just need to be known and trusted and also have an identity card”* (**trekker**)

The findings show that formal training was not a key component in terms of being a trader or a trekker. A key informant at Butula market indicated, *“There is no training that one undergoes; they just come and start the business*” **(KII)**.**”**

> The findings also indicate that information on how to identify sick animals used to be communicated to the traders but that no longer happens. A key informant at Amukura market said;
>
> “*We used to call them in the market and give them some basic education on how to identify sick animals, but we don’t do that anymore. Now we just walk and inspect animals and remove the sick ones from the market*” (**KII)**.
>
> **“***I do not know if they undergo any training, but what I know is that anybody can just come and join the trade provided he or she has capital*” (**KII)**.

In terms of learning the trade, apprenticeship on approximation and negotiation for prices, selection of good animals, and identification of sick animals were mentioned by all of the informants. Some of the responses given included:

> “*Before you start this job, you must have someone who is helping you because you cannot know what size of cow is sold for what price. You must have someone to guide you on the prices*” (**IDI)**.
>
> “*There are people who introduced me to the business, I used to walk with them, and I learned from them*” **(IDI)**.
>
> “*There is no formal training; however, people who have experience in the business must teach you how to negotiate prices. The experienced traders also teach you how to select a good animal that will give you good returns*” **(IDI**).

### Knowledge

The study sought to bring out the actors’ knowledge of zoonotic diseases. Therefore, the questions explored if the informants know about zoonotic diseases if they could name some of the zoonotic diseases, their modes of transmission, and some of the symptoms of zoonotic diseases. It was established that vernacular radio stations, seminars, and workshops have contributed to the actors’ knowledge.

### Knowledge of zoonotic diseases

The findings show that some informants know that diseases can come from animals and infect humans. We note fair knowledge among livestock actors on zoonotic diseases demonstrated by the fact that most respondents know that there are livestock diseases; however, not all of them know that diseases that affect humans can originate from animals. The traders showed more knowledge of zoonotic diseases compared to the trekkers. A majority of the informants named foot and mouth disease (FMD), Brucellosis, Lumpy skin disease (LSD), and Anthrax. This could be because they are the most common diseases in the area, as was backed up by the key informant as shown below

> *“The most common diseases in this area are Lumpy skin disease, foot and mouth, black water and anthrax although we have not had an outbreak recently in this market”* **(KII)**

### Knowledge of symptoms

The actors have knowledge of symptoms of livestock diseases in general. They have ways of identifying sick animals even though they don’t necessarily indicate a zoonotic disease infection, the actors identify sick animals by checking if the animal has: dropping ears, a lot of mucus in the nose, diarrhea, blood-stained urine, and low activity levels. This knowledge is shared by both traders and trekkers.

### Knowledge of modes of transmission

The results revealed knowledge of possible transmission of zoonotic diseases through consumption of meat and milk from infected animals, this was the most common among the respondents.

> *“I know diseases that can come from animals to humans, such diseases can infect you if you eat meat from infected animals … (* **IDI**,)

From the study the level of knowledge varies among the different actors, some can name some of the diseases and the modes of transmission and even identify the symptoms. This is consistent with a study in Tanzania (13) that found patchy knowledge and awareness on zoonotic diseases.

### Practices of actors

The actors have the practice of moving animals from one market to another and some of the animals are moved into Kenyan markets from Uganda and vice versa. This can contribute to the spreading of diseases if an infected animal is moved. There are also practices such as culling sick animals for consumption and consumption of dead animals that have been shown to influence the transmission of zoonotic diseases (29). Practices of livestock traders are likely to expose them to pathogens or control the spread of pathogens. Thus these practices are major contributing factors for zoonotic disease infection in people. Consequently, the informants were asked questions to explore some of their practices to control or spread zoonotic diseases. The questions were asked to explore these practices and the reasons behind them engaging in them. The study was informed of some practices that the actors engage in which are embedded in their cultural and religious beliefs. They engage in these practices because they believe that there is influence from a higher power that they cannot challenge or question. Most of these practices are surrounding the way they would dispose of a dead animal. A question was asked on what the actors would do if an animal under their care died.

> *“In our tradition, most people don’t like throwing meat away because you will throw away your luck, so even if you don’t want to eat it, you call people who want and give it out”* **(IDI**,).
>
> “*Some people say that if you bury a cow you have thrown away your luck and therefore a dead cow should be eaten, but some of us who are born again believe that luck comes from God when a cow dies, we bury it so that we can be safe from other diseases*” **(IDI)**.

The study identified circumstances where some actors will eat meat from an animal that has died from an unknown cause because they believe that throwing away meat will bring bad luck to them. The study also shows a common practice among the actors to skin their animals when they die before burying them. This practice was mentioned by most of the respondents in different markets, and the reason for doing this was given as cultural beliefs. If one does not do this, they will never be successful in the business, as burying an animal with the skin is akin to burying all your wealth and luck.

The study findings indicate that there are practices that the actors in the livestock trade have adopted that help in the prevention of the spread of livestock diseases and to an extent spread of zoonotic diseases. This involves activities such as isolation of sick animals, separating animals that have come from the one market and are bound to another market and further the market chairman have also instituted rules that animals exhibiting any signs of sickness is not allowed in the ring where the animals are traded. In some markets, there are isolation crèche/ structures for animals diagnosed at the market gates. The ones that find their way in are removed, and the owners are told to get help and only return them when they are well. This practice is enforced by the market chairman and his team, “youths.” The traders are also vigilant and will report any animal that appears sick in the livestock ring.

> *“I isolate sick animals, and then I call a veterinary doctor to come and help by treating it. I also ensure that dead animals are buried”* **(IDI)**.
>
> *“If I am moving many animals and one of them falls sick, Isolate it and tie it in a nearby homestead. I then inform the owner of the animal, who will then take action of calling a doctor. Then, I continue with the rest of the animals to my destination”* **(IDI)**.

The practice of isolation of sick animals was reiterated and confirmed by key informants.

> *“When we identify a sick animal in the market, we remove it from the market and advise the owner to get treatment for it and only bring it to the market when it is healed”* **(KII)**

Actors identified uptake of animal vaccination to prevent infection and the spread of diseases. The study found that vaccination exercises are mainly done when there is an outbreak in an attempt to prevent the further spread of the disease, and the government brings vaccinations.

There is also the issuance of movement permits by the veterinary office for animals that are being moved from one county to another.

> *“We issue movement permits to control diseases. Animals from an area with notifiable diseases will not be issued with movement permits “***(KII**,)

## Discussions

Like other similar studies on knowledge of zoonotic diseases, the study revealed that the actors in the livestock trade in Busia do not have detailed knowledge of specific zoonotic diseases or details of disease control. Studies in Sub-Saharan Africa (Swai et al., 2013; Abdi et al., 2015) have pointed to limited knowledge among various actors on zoonotic diseases. We, too, established that many of the informants interviewed in this study know about livestock diseases but not much about zoonotic diseases. Several informants mentioned brucellosis, which shows that some informants know brucellosis as a zoonotic disease. This is consistent with a study by Seyoun *et al*. 2016 (24) on knowledge attitude and practice among small-scale dairy farmers on milk-borne zoonotic diseases. Many study participants know about the potential health risk of drinking raw milk and link this practice to brucellosis. Anthrax, foot, and mouth disease, and Lumpy skin diseases were the diseases that most of the informants mentioned as the most common, and this was also affirmed by the key informants, although there was no recent outbreak of cases reported in the area at the time of the study the fact that most of the actors mentioned these diseases could be attributed to the fact that whenever there is an outbreak, then livestock markets are closed, and quarantine of livestock is enforced. This means there will be no business for them, which could be why most of them mentioned these diseases as common. The traders are also involved in cross-border trade with Uganda, the traders believe that the animals from Uganda are more vulnerable to diseases as compared to the ones from Kenya. The animals from Uganda are therefore sold to butchers for meat and not to farmers for breeding. This could be a considerable risk if indeed the animals have diseases because it will expose the consumers to the risk of infection.

The study revealed that the actors have ways of identifying sick animals. These are not necessarily animals suffering from Zoonotic infections but just ways of knowing if an animal is sick. This is consistent with the findings of Onono *et al*,2019 These ways of identifying sick animals were noted as familiar to both trekkers and traders. This knowledge of identifying sick animals was found to be indigenous knowledge and passed from one person to another and is not acquired through formal training.

Studies have shown that practices such as herding, residing with livestock, slaughtering, skinning, and consuming meat and milk from ill or dead livestock play a vital role in transmitting zoonotic diseases like Rift valley fever (RVF) to humans (25-27). Cultural practices have been shown to play a significant role in disease outbreaks (31-32) This study found that some actors believe that when an animal dies, it must be skinned before it is disposed of, they also believe that meat cannot be thrown away, and therefore when an animal dies, the meat will be eaten or given to people who want to eat. Such practices are based on their cultural belief are deeply rooted. Such practices significantly expose them to the risk of infection.

Results from the study also show that the actors have practices such as separating sick animals from the rest, not allowing sick animals in the market, and vaccination of their animals which can prevent the animals from spreading diseases (34)

According to WHO 2018 (28), there are globally relevant zoonoses that everyone worries about, and locally relevant ones (6) that are important. These diseases can spread rapidly in a particular region (epidemics) or spread widely in many countries worldwide (pandemics), leading to massive losses of life and livelihoods and having a significant economic impact. When compounded with risky beliefs and practices they can quickly wipe out much of the country’s development. This shows that understanding the knowledge attitudes and practices on zoonotic diseases of a community is very important because it provides critical information to help design appropriate control and intervention measures for a zoonotic disease outbreak.

## Conclusions and recommendation

The study revealed patchy awareness and poor knowledge of zoonoses – and livestock health issues more generally – by actors in the livestock trade. Even though the knowledge of zoonotic diseases is low, some of the actors’ practices show that they have some awareness and may take rudimentary measures against them. As shown in other studies, low awareness and poor knowledge of zoonoses combined with food consumption habits in pastoral and agro-pastoral communities are likely to expose the actors to an increased risk of contracting zoonoses. (22) The findings indicate that skinning dead animals and eating carcasses from dead animals is common. It is also evident from the study that the actors (both traders and trekkers) move their livestock from one market to another and move from one county to another, that is Busia, Bungoma, Kakamega, and some of them source animals from Uganda. This practice of the movement of livestock can contribute to the spread of zoonotic diseases from one region to another and across international borders. There is some control to ensure animals don’t move from regions with notifiable diseases like foot and mouth disease or anthrax. This is in the form of issuance of movement permits; however, the study noted that this was not effective as movement permits are only issued in some markets; the study found out that of the markets under study, only one market had movement permits being issued and it was only done to animals that had been sold. There was no one checking if the animals coming to the market were from regions free of diseases. Even though a number of the actors in the livestock trade interviewed do not know zoonotic diseases, they have in place practices that can help control these diseases like vaccination of animals, washing animals, separating animals for sale and those being kept at home, and also ensuring that sick animals do not enter the market. These practices are likely to reduce the risk of animals getting infected and, in turn, infecting them with zoonotic diseases.

Emanating from the above findings, it is evident that Cultural issues are important considerations in the control of infectious diseases hence the importance of incorporating cultural epidemiology in the development of disease prevention, management, and control programs or interventions. The study finds that the locals have a high propensity to information from vernacular radio stations, seminars and workshops therefore it recommends the use of these for awareness creation on zoonotic diseases. There is a need for cross-border intervention programs such as education and sensitization of the actors in livestock trade in this Lake Victoria Crescent ecosystems to help strengthen disease surveillance and control.

## Data Availability

All the data is available and can be requested through the my email address

## Acknowledgments

We thank all the livestock traders and trekkers who participated in the study. A special thanks to Fracoline Awuori, who assisted with data collection. This work was supported by the Biotechnology and Biological Sciences Research Council, the Department for International Development, the Medical Research Council, the Natural Environment Research Council, and the Defence Science & Technology Laboratory, under the Zoonoses and Emerging Livestock Systems (ZELS) program, grant reference BB/L019019/1, and the CGIAR Agriculture for Nutrition and Health (A4NH) program, led by the International Food Policy Research Institute (IFPRI). We also acknowledge the CGIAR Fund Donors (hppt://www.cgiar.org/funders/).

